# Disturbed respiratory viral ecology by COVID-19 pandemic reveals the pivotal role of Influenza in virus-virus interaction network

**DOI:** 10.1101/2025.01.11.24319626

**Authors:** Simeng Liang, Jianbo Xia, Chunchen Wu, Yongliang Zhao, Xiyu Lan, Yanran Li, Xingyu Nan, Liang Qu, Chanjuan Zhou, Yuanan Zhao, Shi Liu, Chao Shen, Zishu Pan, Mingzhou Chen, Ke Lan, Ke Xu

**Affiliations:** State Key Laboratory of Virology and and Biosafety, Taikang Center for Life and Medical Sciences, College of Life Sciences, Wuhan University, Wuhan 430072, P.R. China; Department of Laboratory Medicine, Maternal and Child Health Hospital of Hubei Province, Tongji Medical College, Huazhong University of Science and Technology, Wuhan 430070, China; Hubei Jiangxia Laboratory, Wuhan, 430200, China; Institute for Vaccine Research, Animal Biosafety Level 3 Laboratory, Wuhan University, Wuhan 430072, Hubei, P.R. China; Frontier Science Center for lmmunology and Metabolism, Wuhan University, Wuhan 430072, Hubei, P.R. china

## Abstract

Concurrent epidemics of respiratory viruses provide avenues for intricate virus-virus interactions, yet how molecular-level viral interactions patterns shape viral ecology and epidemic dynamics remain enigmatic. Here, we present real-world virus crosstalk by comprehensively analyzing diagnostic data from a large cohort (37,415 respiratory illness cases pre-COVID-19 pandemic, 22,239 cases thereafter), mainly infants/toddlers, sourced from the same local hospital. Such high-risk group cohort allowed us to examine consistent coinfections among 7 respiratory viruses, despite under an overall reduced infection rates due to COVID-19 disruption. We explored drivers of stable ecosystem and identified a directional virus-virus interaction network between influenza and other respiratory viruses. Monthly prevalence patterns analysis of individual virus revealed IAV positively interacted with RSV, characterized by synchronous seasonality (ρ= 0.67). Conversely, IAV negatively interacted with HPIV3, marked by asynchronous seasonality (ρ= -0.56). Sequential/simultaneous coinfection experiments further confirmed two viruses could contact in the same cell but show distinct coinfection outcomes, such as IAV significantly augmented RSV infection but inhibited HPIV3. We further demonstrated coinfection with IAV and RSV led to exacerbated lung damage in mice, while were associated with aggravated disease outcomes among children. Post-COVID-19, we observed a notable suppression in the spread of respiratory viruses, with a particularly sharp decline in influenza. This reduced influenza activity disrupted virus interactions between influenza and other respiratory viruses, driving the concurrent resurgence of other respiratory viruses. When influenza gradually returns to circulation, the interactions could be reinstated, shaping respiratory virus circulations in a predictable and typical pattern. These findings underscore the pivotal role of influenza in directional interplays among respiratory viruses that shape viral ecology.

**Striking image:** 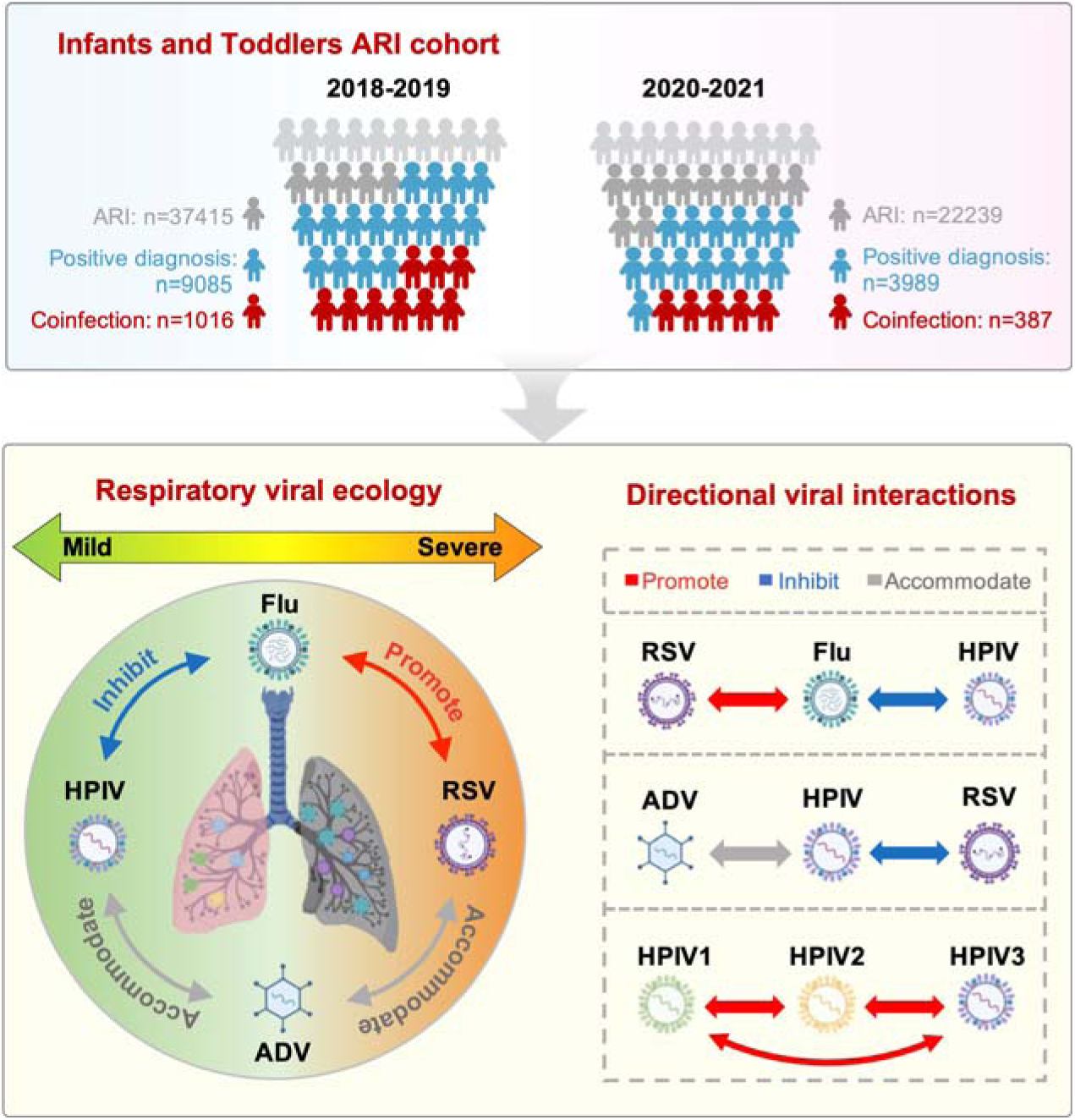

**The respiratory viral ecology with directional virus-virus interactions.:** A substantial cohort of infants and toddlers, comprising 37,415 cases of acute respiratory infection (ARI) pre-COVID-19 pandemic and 22,239 cases thereafter, underwent testing for seven human respiratory viruses: ADV, RSV, IAV, IBV, HPIV1, 2, and 3. The prevalence of coinfection with at least two viruses was 11.18% (pre-) and 9.70% (post-) respectively, showcasing a stable and intricate ecosystem of multiple respiratory viruses even amidst the global disruption caused by COVID-19. Findings from experimental coinfections are consistent with viral seasonal dynamics, where positive interactions (red arrows), such as Flu promoting RSV, exhibit synchronous seasonal patterns, whereas negative interactions (blue arrows), like Flu inhibiting HPIV, display asynchronous seasonal trends. Viruses that demonstrate no interactions with each other (gray arrows), like ADV and RSV, can coexist harmoniously within the host environment (accommodate).

## Introduction

The COVID-19 pandemic has raised significant concerns regarding respiratory viruses. In addition to SARS-CoV-2, a range of respiratory pathogens, including influenza virus, respiratory syncytial virus (RSV), parainfluenza virus (HPIV), and others, occupy overlapping ecological niches in the respiratory tract, thereby creating favorable conditions for coinfections^1–3^. Epidemiological studies have consistently documented the prevalence of respiratory virus coinfections in individuals, with rates exceeding 10%^3–7^, or even reaching 42.7% in some cases^8^. Despite the high coinfection occurrence of respiratory viruses observed in nature, the intricate molecular interactions between these viruses and how COVID-19 pandemic modulate these interactions have yet to be fully elucidated.

Mathematical models have been proposed to evaluate virus interactions based on clinical diagnostic data, yet these investigations have yielded inconclusive and conflicting findings^9–13^. For instance, one study indicated a positive interaction between human common coronavirus (HCoV) and HPIV3^11^, while another analysis proposed a negative interaction between them^12^. Different studies have reported conflicting results in influenza viruses and RSV^13,14^, with one showing a negative interaction^14^ and the other suggesting no interference between the two viruses^13^. These discrepancies may arise from variations in clinical sources, differing local climate conditions, and diverse mathematical models used. Additionally, patient age is a key determinant for shaping viral coinfection patterns and was not uniform in many studies, as the coinfection rate in children is substantially higher than in adults or the elderly^11,15^. Therefore, virus-virus interactions built on mathematic models of clinical diagnostic data have limitations from case to case, which need to be validated with experimental evidence.

Our previous study found that IAV promoted SARS-CoV-2 infectivity in co-occurring cell cultures, ultimately exacerbating lung damage in coinfected mice^16^. This experimental positive interaction between IAV and SARS-CoV-2 aligned with a high coinfection rate of IAV and SARS-CoV-2 observed in clinical respiratory illness^17–20^. Another study demonstrated an experimental negative interaction between IAV and rhinovirus, potentially explaining the interruption of the 2009 influenza pandemic by the annual autumn rhinovirus epidemic^21^. These findings underscored that virus-virus interaction at a molecular level is linked to the interconnected epidemiology patterns of viral infections in clinical observations.

In this study, we combined statistical and experimental analysis to elucidate the intricate viral interactions among seven respiratory viruses. Spearman analysis of monthly prevalence time series for each pair of respiratory viruses unveiled robust correlations, particularly between influenza virus and non-influenza viruses, such as a positive correlation between IAV and RSV (ρ= 0.67). Consistent with the result of Spearman analysis, experimental evidence proved that IAV promoted RSV infection in co-cultured cells. These findings suggested that virus-virus interactions are determinant factors for virus epidemic patterns. During the post-COVID-19 outbreak period from 2020 to 2021, influenza activity significantly declined. The disrupted virus-virus interactions between influenza and non-influenza viruses resulted in irregular seasonal patterns for RSV and other respiratory viruses. Since influenza has resurged in the winter season of 2023, wherein the interactions between influenza and non-influenza viruses could be reinstated, shaping respiratory virus circulations in a predictable and typical pattern.

## Results

### Alterations in seasonal prevalence patterns of respiratory viruses attributed to the COVID-19 pandemic

To explore the impact of the COVID-19 pandemic on other respiratory viruses, we analyzed the diagnostic data from respiratory illness cases, mainly infants/toddlers (87.7% aged 0-4) sourced from the same local hospital, tested with a seven-virus multiplex immunofluorescence panel, encompassing 37,415 cases pre-COVID (2018-2019) and 22,239 post-COVID (2020 to 2021) (Table 1). Pre-COVID, the detection rate with at least one virus stood at 24.28% (9085/37,415), which was comparable to previous studies^11,13,21^. However, subsequent to the pandemic, this rate observed a slight decline, dropping to 17.94% (3989/22,239), suggesting potential alterations in viral circulation patterns due to the pandemic’s disruption (Table 1). We therefore evaluated monthly prevalence patterns across all the seven viruses (Figure 1). The results revealed a distinct seasonal prevalence pattern for respiratory viruses pre-COVID. Specifically, influenza viruses (depicted in red) and RSV (represented by yellow) exhibited peak prevalence during the winter months, whereas the HPIV family (illustrated in blue) reached its peak prevalence during the summer. Notably, ADV (dark purple) demonstrated a prevalent presence throughout the year, underscoring its persistence and lack of pronounced seasonal variation (Fig. 1a). In the merged curve analysis, IAV and RSV exhibited a synchronous seasonal cocirculation pattern, while IAV and HPIV3 displayed an asynchronous cocirculation trend (Fig. 1b). Furthermore, HPIV family, HPIV1, HPIV2, and HPIV3, shared similar seasonal patterns, with HPIV-3 emerging as the dominant virus among them (Fig. 1b). Post-COVID, seasonal patterns disrupted, notably influenza’s decline (Fig. 1c), triggering a surge of non-influenza viruses losing their seasonal trends (Fig. 1d). Notably, HPIV family showed an unusual winter peak in 2020, highlighting atypical patterns.

**Fig. 1.**
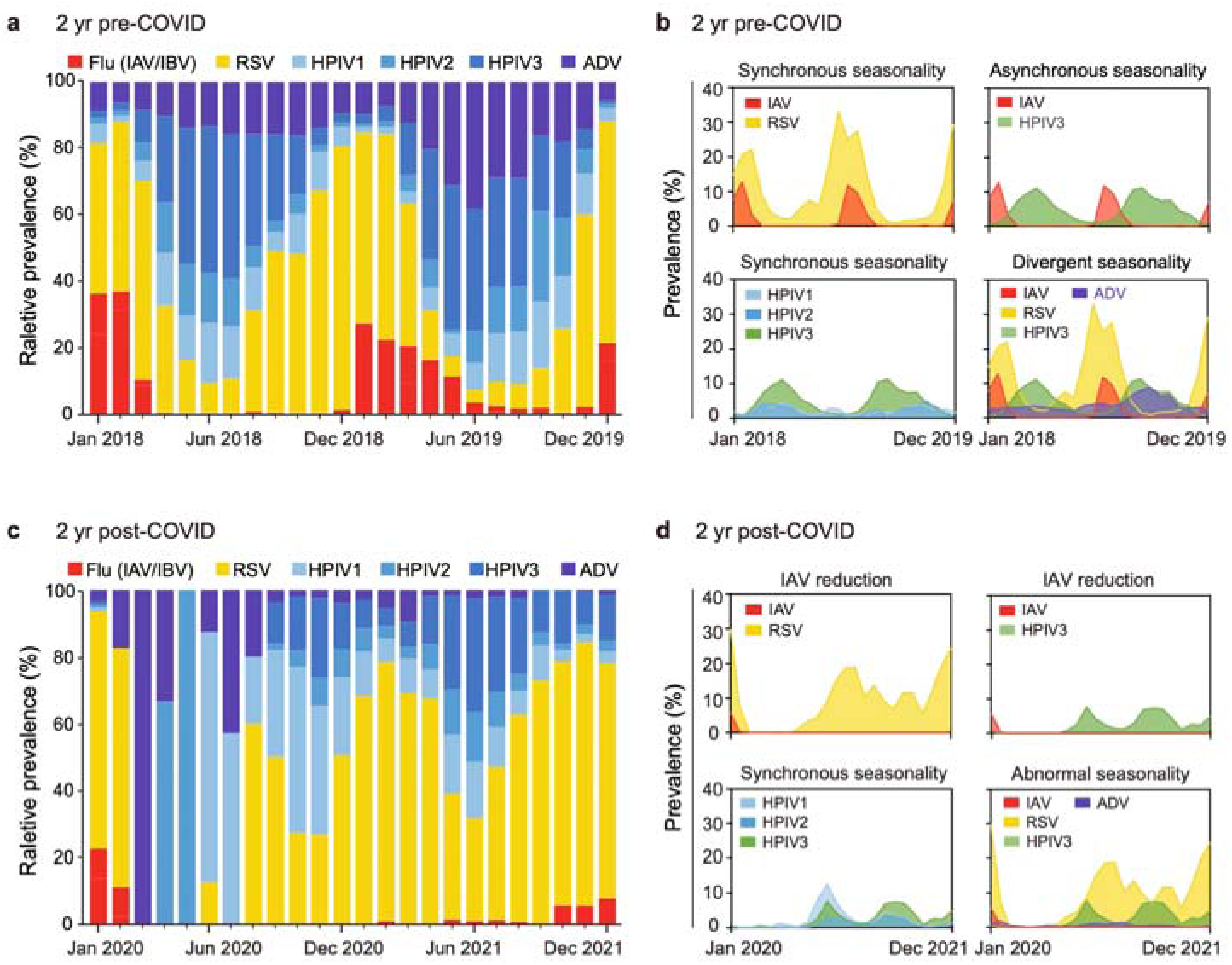
Seasonal prevalence patterns of viral respiratory infections from 2018 to 2021. Prevalence was measured as the proportion of patients testing positive for a given virus among those with positive diagnoses in each month. Jan = January, Jun = June, Dec = December. **a**, Relative virus prevalences in each calendar month from 2018 to 2019 (2 yr pre-COVID). Note total virus counts may sum to more than those informing single infection prevalences due to coinfections. **b**, Comparative prevalences of viral infections showing synchronous or asynchronous seasonality. **c**, Relative virus prevalences in each calendar month from 2020 to 2021 (post-COVID). **d**, Comparative prevalences of viral infections detected among patients post-COVID. **a**-**d**, Flu = influenza virus, including IAV = influenza A virus and IBV = influenza B virus; RSV = respiratory syncytial virus; HPIV1 = human parainfluenza virus 1; HPIV2 = human parainfluenza virus 2; HPIV3 = human parainfluenza virus 3; ADV = human adenoviruses.

**Table 1.** Positive rates of viral pathogens among patients with acute respiratory infection (ARI) in HuBei Province of China, pre- and post-COVID.

|  | **Pre-COVID** |  | **Post-COVID** |  |
| --- | --- | --- | --- | --- |
|  | Total cases (N) | Positive diagnosis (N,%) | Total cases (N) | Positive diagnosis (N,%) |
|  | 37415 | 9085 (24.28) | 22239 | 3989 (17.94) |
| Sex N (%) |  |  |  |  |
| Male | 22299 (59.6) | 5495 (60.48) | 13162 (59.2) | 2401 (60.19) |
| Female | 15116 (40.4) | 3590 (39.52) | 9077 (40.8) | 1588 (39.81) |
| Age N (%) |  |  |  |  |
| <6 month | 6938 (18.5) | 1924 (21.18) | 3873 (17.4) | 708 (17.75) |
| 6-11 month | 5764 (15.4) | 1764 (19.42) | 2992 (13.5) | 683 (17.12) |
| 1-4 year | 20073 (53.6) | 4746 (52.24) | 12663 (56.9) | 2466 (61.82) |
| 5-17 year | 4640 (12.4) | 651 (7.17) | 2711 (12.2) | 132 (3.31) |

### Persistent coinfection patterns among respiratory viruses

To investigate whether virus-virus interactions contributed to altered seasonal prevalence patterns during the COVID-19 pandemic, a detailed analysis of mono-infections and mixed infections within this large cohort was conducted. The introduction of SARS-CoV-2 into the ecosystem led to a general decline in infections across all seven viruses examined (Figs. 2a-g). Among them, ADV was the most significantly impacted, with cases dropping from 1494 pre-COVID to 139 post-, with a 6.33-fold reduction in positivity (3.99% to 0.63%). IAV was the next most impacted, exhibiting a 5.85-fold decrease in cases (from 897 to 92) (Fig. 2b). The number of IBV infections declined from 367 to 111, resulting in a 1.96-fold reduction in positivity (Fig. 2c). Similarly, HPIV3 cases decreased from 1860 to 780, with a 1.42-fold decrease in positivity (Fig. 2d). HPIV2 infections also dropped, from 755 to 334, accompanied by a 1.35-fold reduction in positivity (Fig. 2e). In contrast, while the number of HPIV1 cases declined from 890 to 708, the positivity rate slightly increased by 1.34-fold (2.38% to 3.18%) (Fig. 2f). RSV cases decreased from 4355 to 2513, but the positivity rate remained relatively stable, with only a marginal 1.01-fold decrease (11.45% to 11.30%) (Fig. 2g).

**Fig. 2.**
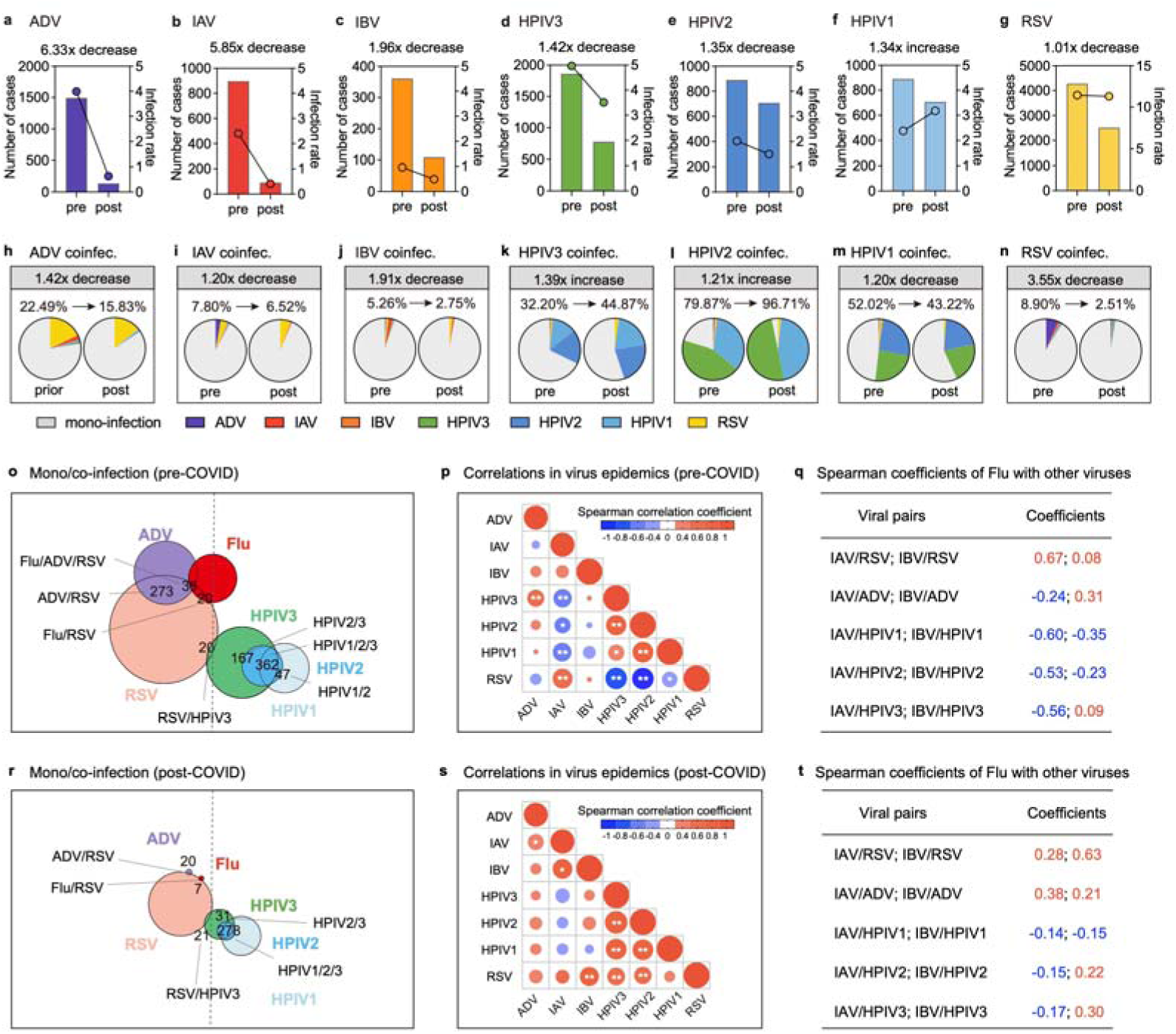
Infection patterns of patients with respiratory illness from 2018 to 2021. **a-g**, Number of infection cases for all seven viral pathogens tested pre- and post-COVID, with fold changes in the infection rate displayed. (**a**) ADV, (**b**) IAV, (**c**) IBV, (**d**) HPIV3, (**e**) HPIV2, (**f**) HPIV1, and (**g**) RSV. **h-n**, Coinfection rates of the seven viral pathogens pre- and post-COVID, illustrated with colored parts in the chart and corresponding fold changes. The colored parts in the chart present the proportion of coinfection with each pathogen for the virus, (**h**) ADV, (**i**) IAV, (**j**) IBV, (**k**) HPIV3, (**l**) HPIV2, (**m**) HPIV1, and (**n**) RSV. **o**, Number of cases (pre-COVID) were clustered by virus pairs (less than 10 cases were omitted). **p**, **q**, Negative and positive correlations analyzed from Spearman’s rank method in monthly viral infection prevalences pre-COVID are shown in blue and red cycles, respectively. The size of points reflects the strength of the correlations. Significant correlations are indicated by *, ρ > 0.406; **, ρ > 0.521. **q**, The correlation coefficients from Spearman analysis of viral infection time series between IAV or IBV and other viruses were shown. **r**, Number of cases (post-COVID were clustered by virus pairs (less than 7 cases were omitted). **s, t**, Negative and positive correlations analyzed from Spearman’s rank method in monthly viral infection prevalences post-COVID. **t**, The correlation coefficients from Spearman analysis of viral infection time series between IAV or IBV and other viruses were shown.

Recognizing infants and toddlers as susceptible hosts for mixed infections, we delved into an analysis of viral coinfections (Figs. 2h-n). Pre-COVID, 11.18% (1016/9085) of patients were found to be infected with at least two viruses. Interestingly, despite a decrease in the absolute number of coinfection cases to 387 post-pandemic, the coinfection rate remained fairly constant, constituting 9.70% (387/3989) of cases. Pre-COVID, 22.49% of ADV-positive patients had coinfections, with RSV predominating (79.95%). Post-pandemic, 15.83% of ADV-positive patients still had coinfections, with RSV still the most frequent coinfecting virus (90.91%). Notably, the ADV coinfection rate decreased modestly by 1.42-fold (Fig. 2h), significantly less than the 6.39-fold drop in ADV positivity (Fig. 2a). Among IAV-positive patients, 7.80% had coinfections pre-COVID (RSV and ADV top, 45.22% and 36.65% respectively). Post-pandemic, 6.52% remained coinfected (RSV predominant, 83.33%). The IAV coinfection rate declined slightly by 1.20-fold (Fig. 2i), less than the 5.80-fold drop in positivity (Fig. 2b). For IBV, coinfection dropped from 5.26% to 2.75%, similar patterns, with the decrease in coinfection rate comparable to that in positivity (Fig. 2j). HPIV3 (Fig. 2k) and HPIV2 (Fig. 2l) coinfection rates increased by 1.39-fold and 1.21-fold respectively, while HPIV1 (Fig. 2m) barely decreased by 1.20-fold; mainly within HPIV families. RSV coinfection declined from 8.90% to 2.51% post-pandemic (Fig. 2n), largely due to the significant drop in ADV positivity.

### Paired correlation in viral epidemic patterns

To uncover the underlying factors sustaining stable coinfection patterns, we visualized the number of infection cases by clustering virus pairs. Each colored circle represents the number of infection cases for each virus, while the overlapping area between circles indicates cases where patients were concurrently infected with both viruses (Figs. 2o and 2r). Notably, our analysis distinguished two distinct clusters: one comprising ADV, Flu, and RSV, and the other encompassing HPIV. We further explored the dynamics between respiratory viruses utilizing Spearman correlation analysis on monthly prevalence data. The correlation strength was visualized by point size (Figs. 2p and 2s). Pre-COVID (Fig.2p), we found strong positive correlations between HPIV1 & HPIV2 (ρ= 0.76) and IAV & RSV (ρ= 0.67, red), as well as top negative correlations between RSV & HPIV3 (ρ= -0.74) and IAV & HPIV3 (ρ= -0.56, blue). However, post-COVID (Fig. 2s), there were observable weakenings in both positive and negative interactions, with an overall trend of positive correlation shift, suggesting a synchronized resurgence of respiratory viruses following the pandemic. Intriguingly, the Spearman correlation coefficients of Flu (IAV or IBV) with other respiratory viruses reveal statistically significant patterns predominantly observed pre-COVID (Fig. 2q). Specifically, RSV and ADV demonstrate positive correlations with Flu, whereas the HPIV family consistently exhibits negative correlations. This suggests that the reduction in Flu cases may have contributed to shaping atypical epidemic patterns of these viruses post the pandemic (Fig. 2t), highlighting the dynamic interplay among respiratory viruses.

### Directional virus-virus interplay between influenza and non-influenza viruses in co-occurred cells

To verify molecular-level viral interactions, we developed mono-infection and sequential coinfection models in A549 cells (Fig. 3a). In sequential coinfection, influence of virus A on subsequent virus B infection was analyzed by infecting cells with virus A for 12h, then virus B for 24h. Mono-infected controls (36h A or 24h B) served as benchmarks. Viral load ratios in coinfections, normalized against their respective mono-infection counterparts are shown in Figs. 3b-g. The data in Fig. 3b showed pre-infection with IAV significantly augmented RSV infectivity, yielding a 64.53-fold surge in RSV titer compared to mono-infection. Conversely, IAV had no discernible impact on ADV infectivity but potently suppressed the infectivity of IBV (by 21.50-fold), HPIV2 (by 81.29-fold), and HPIV3 (by 161.61-fold). In the context of IBV, RSV infectivity was marginally enhanced by 2.17-fold, whereas IBV inhibited the infectivity of IAV (by 3.59-fold), HPIV2 (by 34.85-fold), and HPIV3 (by 58.77-fold). Notably, among non-influenza viruses such as RSV (Fig. 3d), HPIV2 (Fig. 3e), HPIV3 (Fig. 3f), and ADV (Fig. 3g), negative or negligible interactions were predominantly observed.

**Fig. 3.**
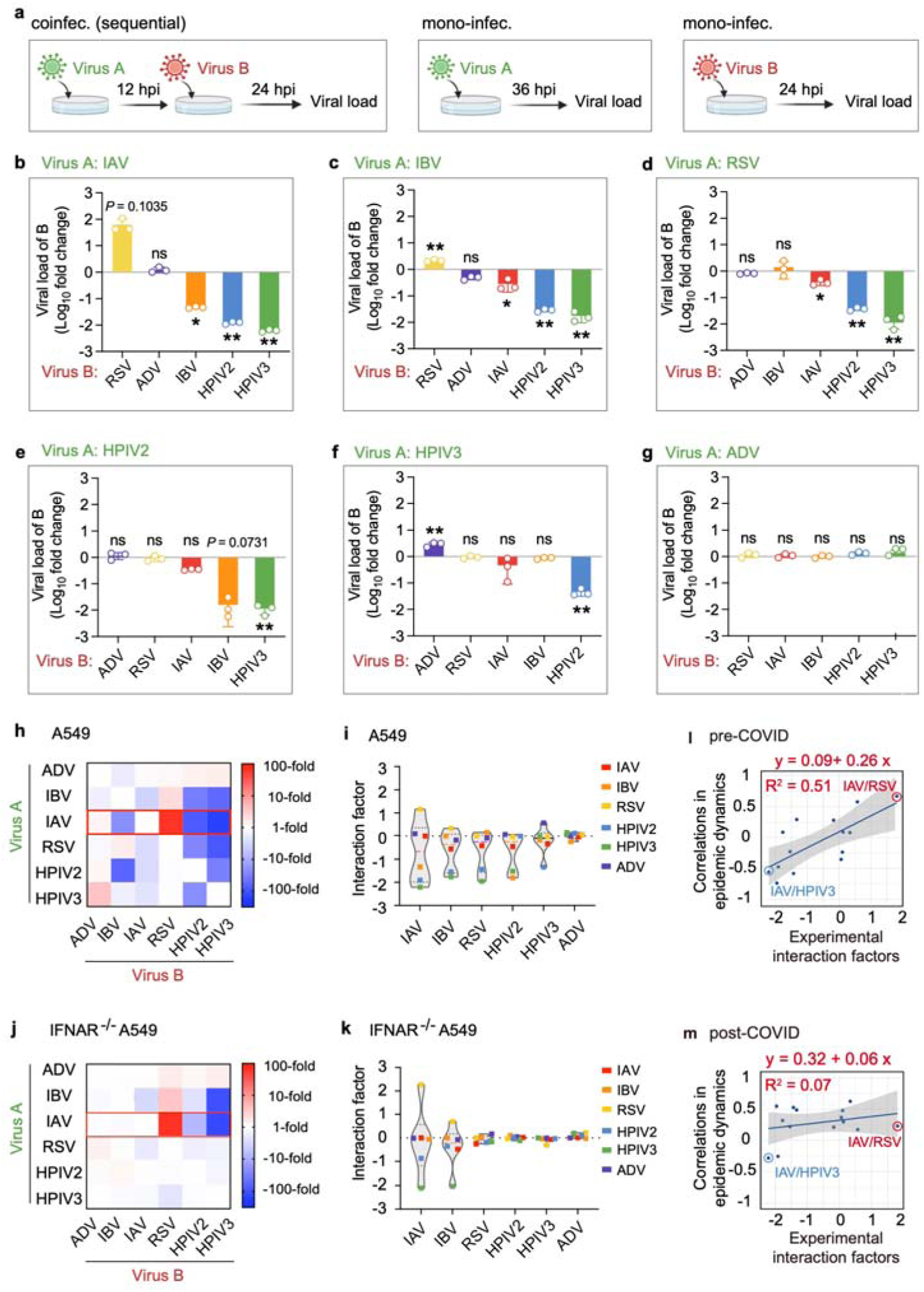
Effect of primary virus infection on secondary virus infection in co-occurring cell cultures. **a**, Experimental procedure diagram for mono-infections and sequential coinfections. For mono-infections, A549 cells were infected with virus A (MOI=0.1) for 36 h (except for adenovirus, which required 60 h) or virus B (MOI=0.1) for 24 h (adenovirus infection took 48 h). For coinfections, cells were initially infected with the primary virus (virus A) at an MOI of 0.1. At 12 hpi, cells were subsequently infected with the secondary virus (virus B) for an additional 24 h (adenovirus infection took 48 h). The viral load in cell lysate was assessed using reverse-transcription quantitative PCR (RT-qPCR) to reflect virus infectivity. **b-g**, The viral load was expressed as fold changes in virus genome copy number in coinfections compared to mono-infections, under pre-infection with (**b**) IAV, (**c**) IBV, (**d**) RSV, (**e**) HPIV2, (**f**) HPIV3, (**g**) ADV. Up panel: Viral load of B; Lower panel: Viral load of A. Values represent means ± SD from three independent experiments. *P < 0.05, **P < 0.01. **h**, **i**, The effect of the primary virus (Virus A) infection on the secondary virus (Virus B) in coinfections. The data was indicated as the mean fold changes of the Virus B viral load in coinfection compared to Virus B mono-infections from three independent experiments in A549 cells. (**h**) Blue and red represent decreased and increased viral load in coinfections compared to the Virus B mono-infections. Red rectangle: the effect of IAV on other respiratory virus infections. (**i**) For the interaction factor, the logarithm with base 10 (log_10_) of the mean fold change was calculated. **j**, **k**, The effect of the primary virus infection on the secondary virus in IFNAR^−/−^ A549 coinfected cells was shown. **l**, Relation between correlations in viral epidemic dynamics and viral interactions fitted by linear regression analysis. The x-axis shows viral interaction factors verified in Fig. 3i, and the y-axis shows the correlations in viral epidemical dynamics evaluated by Spearman analysis in Fig. 2i. **m**, Relation between viral epidemic dynamics post-COVID and viral interaction factors fitted by linear regression analysis.

The above data reveal that the majority of viral interactions are inhibitory, with IAV serving as a notable exception, capable of eliciting both positive and negative crosstalk within the viral network (evident in Fig. 3h’s heatmap and Fig. 3i’s violin plot). The prevalent negative interactions lead us to hypothesize that the host’s innate immune response, particularly the interferon (IFN) response, induced by the primary viral infection (Virus A) may adversely impact subsequent Virus B infection. We therefore replicated our mono- and co-infection experiments in IFNAR^-/-^ A549 cells. As shown in Figures 3j and 3k, when the IFN response was abolished, most negative interactions dissipated, and pre-infection with IAV potently enhanced RSV infectivity by 184.70-fold, exceeding the enhancement observed in IFN-competent cells (Fig. 3i). Again, the inhibition of HPIV3 by IAV intensified to 128.68-fold, albeit slightly weaker than in IFN-competent cells. Conversely, coinfections involving RSV, HPIV2, HPIV3, and ADV exhibited no significant impact on each other’s infection in IFNAR^-/-^ A549 cells, suggesting their compatibility and potential for coexistence. These findings in all indicate that the influenza virus’s influence on other respiratory viruses is directional and independent of IFN signaling.

To link the clinical correlation and experimental evidence, a linear regression analysis was undertaken. This analysis yielded a statistically significant positive association (R² = 0.51) between viral epidemic correlations (as presented in Fig. 2p) and experimentally observed viral interactions (data depicted in Fig. 3k). Pre-COVID, more robust virus-virus interactions corresponded closely with epidemic dynamics correlations (Fig. 3l). For example, the enhancement of RSV infectivity by IAV (a 64.53-fold increase shown in Fig. 3k) aligns neatly with their positive correlation during epidemic periods (ρ= 0.67 as seen in Fig. 2p). In contrast, the inhibition of HPIV3 by IAV mirrors their asynchronous seasonal patterns. However, post-COVID, the correlations among the seven respiratory viruses largely diminished, leading to a weakened association as illustrated in Fig. 3m (R²= 0.07). These data underscore the potential for molecular-level virus-virus interactions to modulate epidemic dynamics, emphasizing the pivotal role that influenza viruses play within respiratory ecology.

### IAV promotes RSV infection cycle in co-occurred cells

Given the strongest enhancement of RSV infectivity by IAV, we further examined the interplay between these two viruses. First, we replicated the experiment outline in Figure 3, infecting various cell lines (MDCK, 16HBE, WI-38 VA-13, BEAS-2B) sequentially with IAV for 12 hours, followed by RSV for 24 hours (Fig. 4a). Results revealed a universal enhancement of RSV infectivity by IAV, albeit to varying degrees across cell lines (Figs. 4b–e). Monitoring RSV titers over time (2–72 hours post-RSV infection) showed that IAV facilitated both RSV entry and replication (Figs. 4f and 4g). Notably, RSV entry was unaffected during the initial 0–4 hours post-IAV infection but increased significantly 6–10 hours later, suggesting IAV replication drives this effect (Fig. 4h).

**Fig. 4.**
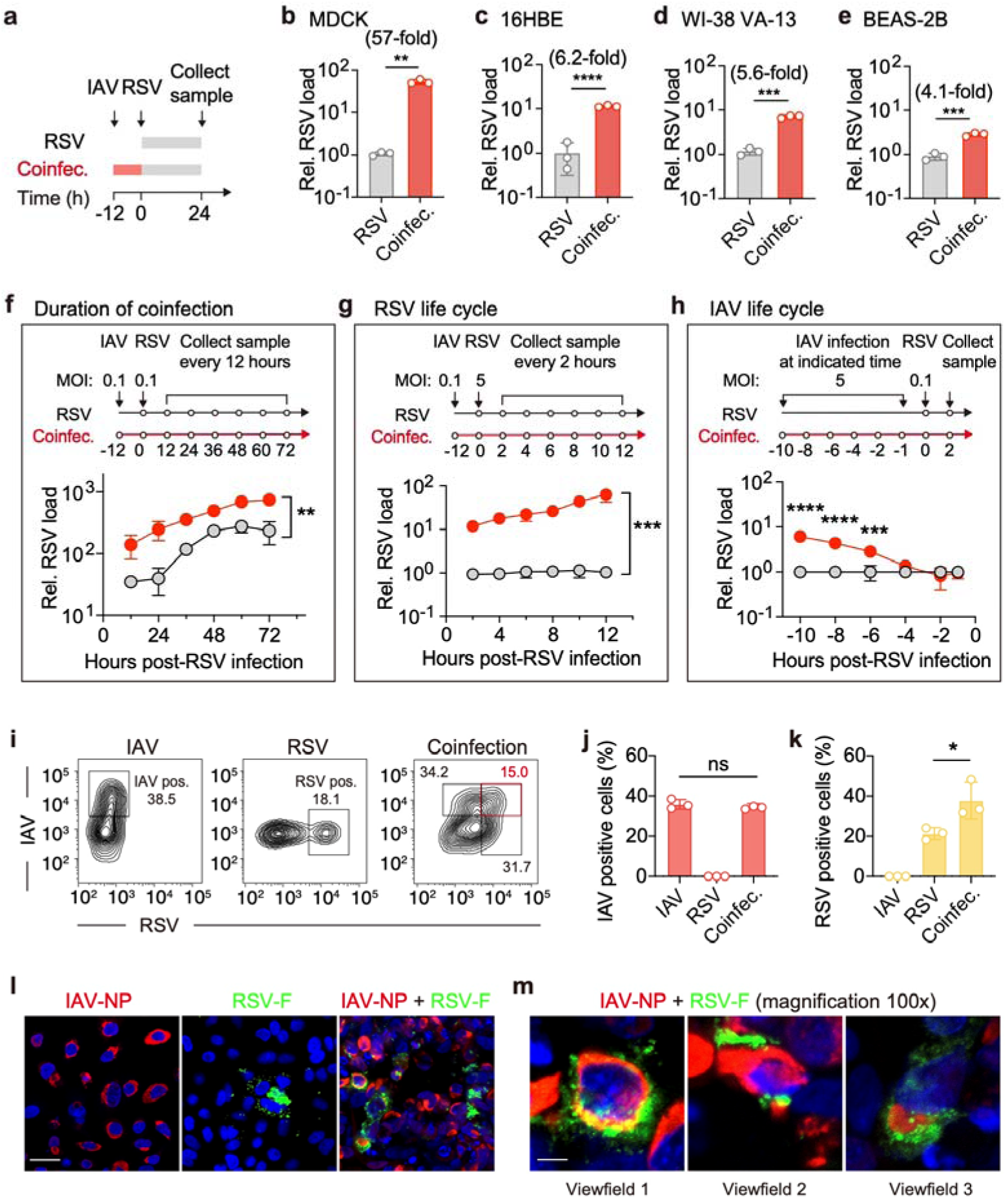
Cellular infection during IAV and RSV coinfections. **a-e**, Cells were initially infected with IAV (MOI=0.1) for 12 h, then cells were subsequently infected with RSV (MOI=0.1) for another 24 h. Coinfections were performed in MDCK(**b**), 16-HBE(**c**), WI-38 VA-13(**d**), and BEAS-2B(**e**) cells. **f**, IAV (MOI of 0.1), RSV (MOI of 0.1). At 12, 24, 36, 48, 60, and 72[h post-RSV infection, relative RSV titers in cell lysates were expressed as fold changes in virus genome copy number in coinfections compared to the mock-infections. **g**, **i,** IAV (MOI of 0.01), RSV (MOI of 1). The proportion of cells with IAV-infected (IAV pos.), RSV-infected (RSV pos.), or IAV and RSV coinfected (Dual pos.) in mono-infections or coinfections at 24 hours post-infection (hpi). **j-k**, Proportion of cells with IAV (**j**) or RSV (**k**)-infected in mono-infections and coinfections. **l-m**, Mono- or co-infected A549 cells were stained by immunofluorescence for IAV nucleoprotein (red) and RSV fusion protein (green) at 36 hpi. Nuclei are stained with DAPI (blue). (**l**), Scale bars indicate 20 μm. (**m**), Data were collected from 3 fields-of-view from each slide in an independent experiment. Scale bars indicate 2 μm. **b**-**h**, **j**-**k**, Values represent means ± SD of three independent experiments. *P < 0.05, **P < 0.01, ***P < 0.001, ****P < 0.0001.

To investigate whether IAV and RSV contact in the same cell or different cells, sequential infections were performed and numbers of individual infected cells were examined via flow cytometry. Coinfection resulted in 15.0% of cells being dual-positive for IAV-NP and RSV-F, indicating simultaneous infection (Fig. 4i, red square). While IAV-positive cell percentages were similar between mono- and coinfection conditions (36.8% vs. 34.1%), RSV-positive cells increased substantially (21.3% vs. 37.8%) (Figs. 4j and 4k). Under confocal microscopy, an obvious portion of cells were positively stained by both anti-IAV-NP and anti-RSV-F antibodies suggesting that the two viruses indeed contacted in one single cell (Figs. 4l and m).

### IAV inhibits HPIV3 infection in co-occurred cells

Next, we tested the negative interactive pair of IAV and HPIV3. IAV inhibited HPIV3 across multiple cell types, with varying levels of suppression (data not shown). This inhibitory effect persisted throughout the 6 to 72-hour period following HPIV3 infection, regardless of the duration of coinfection. Flow cytometry analysis revealed a significant reduction in the proportion of HPIV3-positive cells (anti-HPIV3-F staining) under coinfection conditions (6.96% in the coinfection group vs. 22.52% in the HPIV3-only group). Even if the HPIV3 infection was strongly inhibited by primary IAV infection, there were still 2.90% of cells dual positive for both anti-IAV-NP and anti-HPIV3-F staining. The results suggest that inhibition between the negatively interactive viruses might also occur in the same cell.

### Coinfection with IAV and RSV in mice induces more severe pathology

To assess the impact of coinfection on disease severity, we employed a sequential mono-infection and coinfection model using 6-week-old BALB/c mice, exposing them to IAV or/and RSV. Viral load in infected mice lung tissue and histopathological disease severity were examined at the end of experiments (60h post-sequential infection, Fig. 5a). The coinfected mice showed the most weight loss compared to IAV mono-infected mice (Fig. 5b), although viral loads of either RSV or IAV were not elevated in coinfection (IAV titer even decreased a bit, Fig. 5c). The histological data supported the observation in weight changes that more severe lung pathologic damage appeared in coinfected mice as compared to IAV or RSV mono-infection, characterized by a larger area of alveolar septa thickening surrounded by a mixed inflammatory cell population, pulmonary hemorrhage (Fig. 5d). The changes of the histopathological features in lung tissues were scored based on the severity of three histological criteria and the highest clinical score was assessed in coinfected mice than the mono-infected mice (Fig. 5e). To further assess the IAV and RSV foci in infected lungs, the lung sections were submitted to immune-staining with anti-IAV-NP and anti-RSV-F antibodies. Consistent with coinfected cell cultures, the same lung cells were dual infected by both IAV and RSV (Fig. 5f arrowed).

**Fig. 5.**
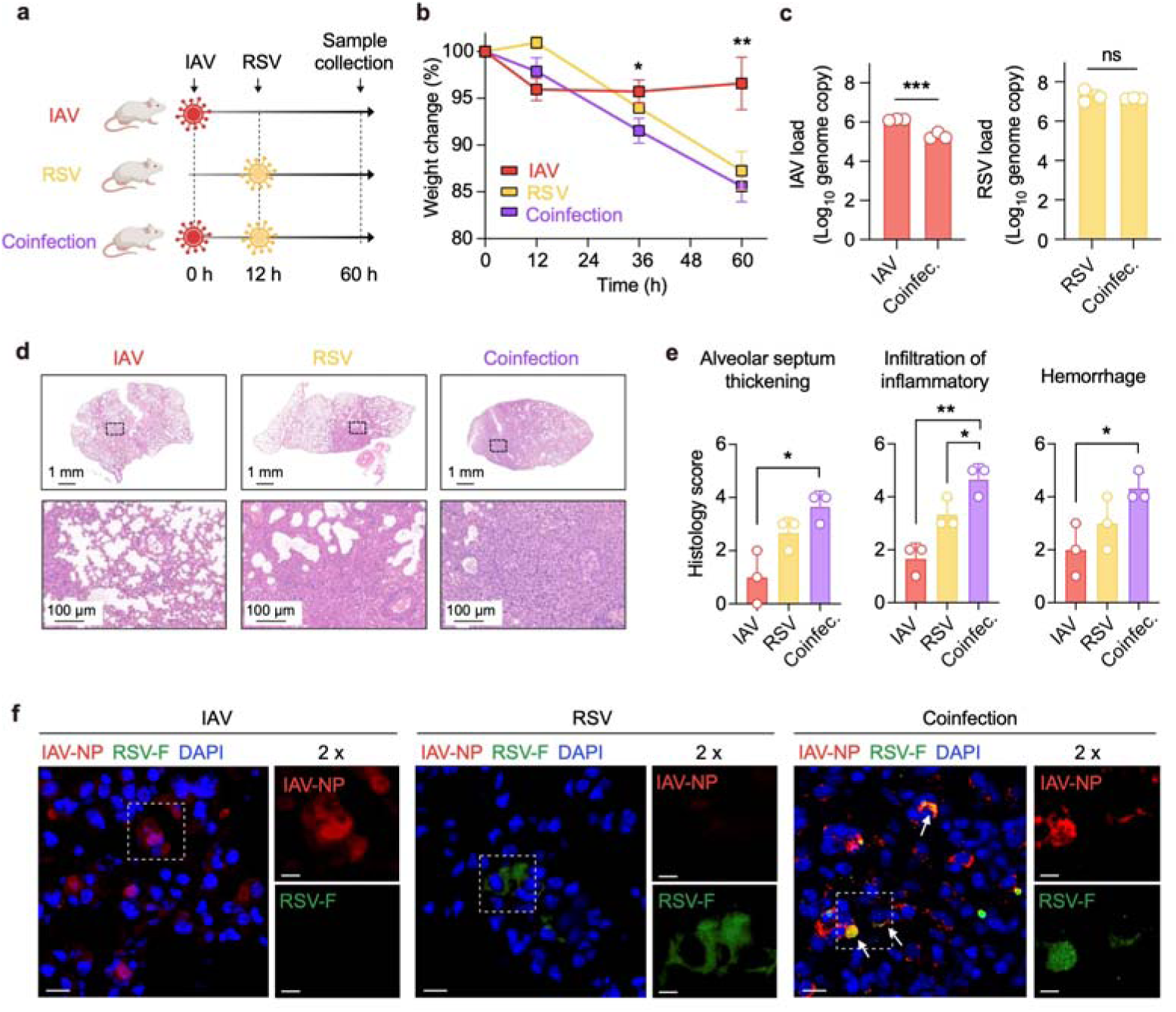
IAV and RSV coinfection induced more severe pathology in infected mice. BALB/c mice were mono-infected or coinfected with IAV or/and RSV sequentially. **a**, Diagram of the experimental procedure. BALB/c mice were initially intranasally infected with 1000 PFU of IAV or PBS. After 12 hours post IAV infection, mice were intranasally infected with 3 × 10^5^ FFU of RSV or PBS. At 60 hours post-IAV infection, lung tissues were collected from all the mice (n=3 in each group). **b**, The body weights and survival were monitored until hour 60. The body weights are presented as the mean percentage of weight change ± SD. The difference in daily weight changes between each group was conducted. **c**, The viral genome copy numbers of IAV (left) and RSV (right) were quantified. Values represent means ± SD of three individual mice. **d**, Histopathologic studies were performed on lung samples from the indicated groups. Scale bars, 1 mm or 100 μm. **e**, Pathological scores were calculated. Values represent means ± SD of three individual mice. **f**, Mono- or co-infected lung cells of mice were fixed and stained by immunofluorescence for IAV nucleoprotein (red) and RSV fusion protein (green). Nuclei are stained with DAPI (blue). Scale bars indicate 20 μm and arrows indicate co-infected cells. White boxes indicate the enlarged regions and scale bars indicate 10 μm.

### Coinfection with Flu and RSV in pediatric patients is associated with aggravated disease outcomes

To evaluate the clinical impact of coinfection, we conducted a retrospective analysis of pediatric medical records, comparing outcomes clinical manifestations of patients with Flu, RSV, or both. The highest coinfection cases pre-COVID occurred in Jan 2019, Mar 2018, Feb 2018, Jan 2018, and Dec 2019 (indicated by arrows in Fig. 6a). From these months, we selected a cohort of 237 Flu, 242 RSV mono-infections, and all 46 coinfections (see Methods for detailed criteria, Fig. 6b). Multiple logistic regression revealed that coinfection, compared to RSV mono-infection, was associated with increased fever (>37.5°C) (Fig. 6d), elevated high-sensitivity C-reactive protein (hs-CRP) levels (Fig. 6e). Conversely, versus IAV mono-infection, coinfection showed varied elevation in pneumonia risk (Fig. 6f), normal lymphocyte maintenance (Fig. 6g), and prolonged hospitalization (Figs. 6h-j).

**Fig. 6.**
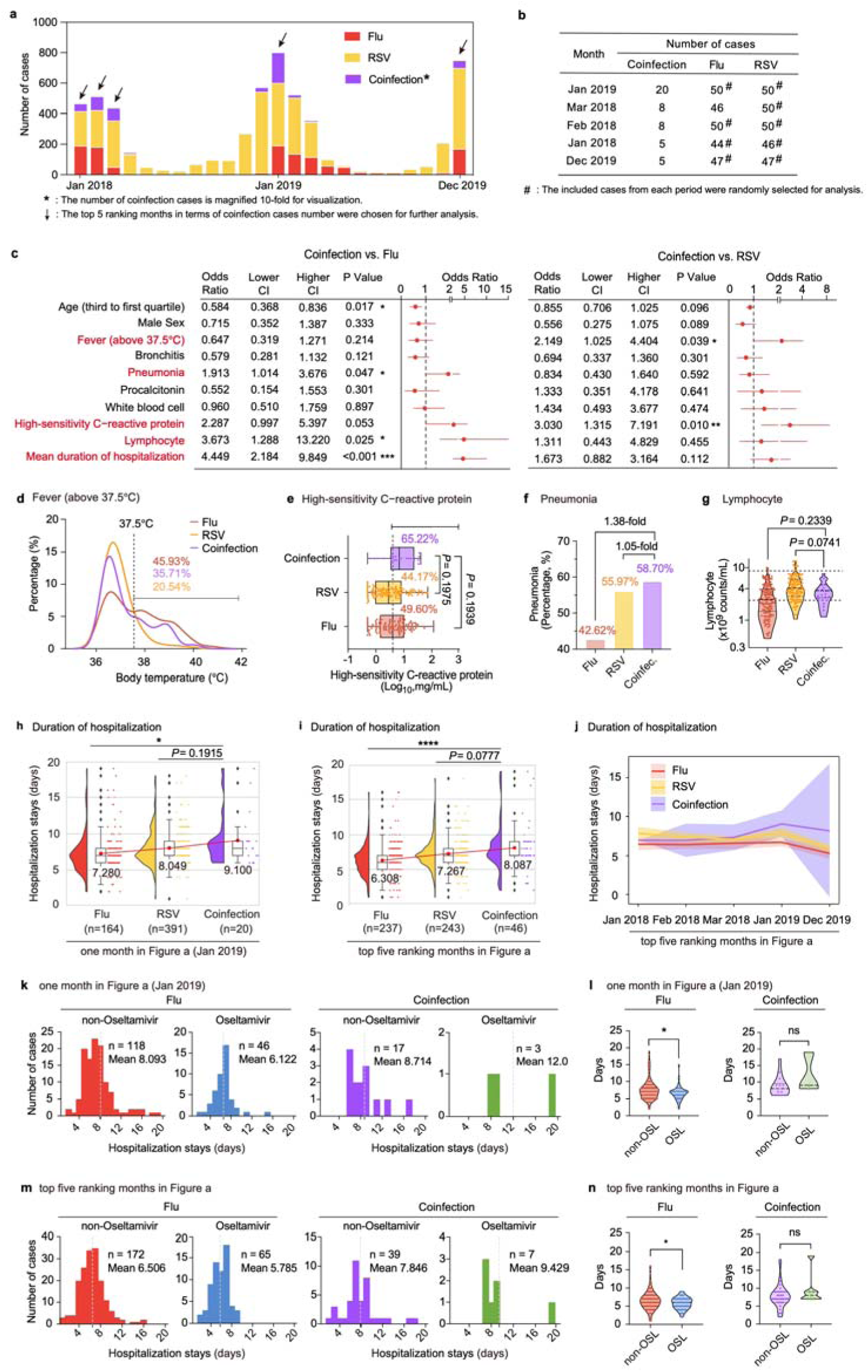
Clinical outcomes of pediatric patients with Flu or/and RSV infection. **a,** The number of Flu or/and RSV infected cases from Jan 2018 to Dec 2019. The number of coinfection cases was magnified 10-fold for visualization. **b**, The number of cases included in our cohort. **c**, Forest plot demonstrating the odds ratio (OR) and 95% confidence interval (CI) for the clinical outcomes with coinfection (vs. Flu or RSV mono-infection) after multivariable logistic regression in our cohort. **d-g**, Comparisons of body temperature (**d**), pneumonia (**e**), high-sensitivity C-reactive protein level (**f**), and lymphocyte count (**g**) between coinfection and mono-infection groups. **h, i**, Duration of hospitalization of cohort in Jan 2019 (**h**) and five indicated months **(i).** Mean hospitalization stays were labeled. **f-i,** *P* values are from unpaired one-way ANOVA. **j,** Mean hospitalization stays ± 95% CI of patients in each month of the five indicated months. **k**, Length of hospitalization duration among the patients infected with Flu alone (left) or coinfected with Flu and RSV (right) enrolled in Jan 2019. The patients were divided into Oseltamivir treatment and non-Oseltamivir treatment group. **l,** Comparison of hospitalization duration of patients enrolled in Jan 2019 with Oseltamivir treatment versus non-Oseltamivir treatment. **m, n,** Comparison of hospitalization duration of patients enrolled in five indicated months with Oseltamivir treatment versus non-Oseltamivir treatment.

To validate these findings, we expanded our analysis to include all Flu and RSV cases from Jan 2019, which had the highest overall coinfection cases. This extended cohort included 164 Flu, 391 RSV mono-infections, and 20 coinfections. Consistently, coinfected patients exhibited significantly longer hospital stays (9.1 days vs. 7.28 days for Flu mono-infections and 8.049 days for RSV mono-infections, Fig. 6h). Given the clinical burden of coinfections, we next investigated the efficacy of existing treatments. Oseltamivir, an anti-influenza drug, was reported to reduce recovery time for patients with influenza-like illness^22,23^. To evaluate its potential benefits for coinfected patients, we analyzed its impact on Flu mono-infections and coinfections. While Oseltamivir significantly reduced hospitalization duration in Flu mono-infected patients, it showed no measurable benefit for coinfected patients (Figs. 6k–n). These findings highlight the need for broad-spectrum antiviral treatments to address the challenges of respiratory virus coinfections.

## Discussion

Multiple co-circulating respiratory viruses occupy overlapping ecological niches in the respiratory tract, creating favorable conditions for coinfections and virus-virus interactions. The currently defined virus-virus correlation is from analyzing clinical diagnostic data, in this study, we combined clinical diagnostic data analysis with experimental infection evidence. We found the implementation of non-pharmaceutical interventions during COVID-19 pandemic indeed limited the spread of respiratory viruses^24^, the infection rate of respiratory viruses in our cohort decreased from 24.28% (9085/37,415) to 17.94% (3989/22,239). Among these viruses, the infection rate of ADV and IAV were largely impacted (ADV, 3.99% to 0.63%, 6.39-fold decreased; IAV, 2.40% to 0.41%, 5.80-fold decreased). Intriguingly, compared with the significantly reduced infection rate, the coinfection rate in ADV- or IAV-positive patients only limited decreased (ADV, 22.49% to 15.83%, 1.42-fold decreased; IAV, 7.80% to 6.52%, 1.20-fold decreased), suggesting a stable virus-virus interaction among these respiratory viruses.

To illustrate the virus-virus interactions, we performed Spearman analysis to evaluate the correlations in the monthly prevalence time series for each pair of respiratory viruses. The results showed the correlations among respiratory viruses existed pre-COVID, but the strength of correlations diminished post-COVID. Pre-COVID, 5 positive interactions and 6 negative interactions were observed among respiratory viruses. The strongest positive correlations existed among HPIV family (HPIV1 and HPIV2, ρ= 0.76; HPIV2 and HPIV3, ρ= 0.73; HPIV1 and HPIV3, ρ= 0.48). Other similar studies did not classify the HPIV subtypes, and thus neglected the positive interactions among the HPIV family^7,25^. Our findings suggest that more attention should be paid to coinfections among the HPIV subtypes.

The second strongest positive correlation is between IAV and RSV (ρ= 0.67). Our clinical analysis was conducted in children (87.6% infants and toddlers, and 12.4% school-age children), whose immature immune systems make them more susceptible to respiratory viruses such as RSV. Another study was conducted in all-age patients and also showed a positive correlation between IAV and RSV (ρ= 0.425). When they applied the Bayesian hierarchical model to adjust the effect of age, the positive correlation between IAV and RSV was attenuated (ρ= -0.13), indicating that the virus interaction is dependent on the age of the infected individuals^13^. Therefore, the positive interaction between IAV and RSV is usually neglected in clinical analysis involving all-age cohorts^11,14^. We also found ADV positively interacted with HPIV3 (ρ= 0.58), which was verified by other study^13^. However, our findings showed this positive interaction was only presented between ADV and HPIV3, rather than HPIV1 or HPIV2, indicating a more significant contribution of HPIV3 in the virus-virus interaction network.

Our experimental data on coinfections provide empirical evidence that positively correlated viruses in epidemic dynamics indeed exhibit an enhanced capacity for viral infection. This may be attributed to the shared infection-related key factors between the positively correlated viruses. One study indicated that HIV promoted HCV replication through the up-regulated expression of transforming growth factor (TGF)-β1^26^. Another study showed that antibodies elicited by dengue virus could facilitate Zika virus infection^27^. Our previous study demonstrated that influenza virus enhanced SARS-CoV-2 infection by upregulating the expression of the SARS-CoV-2 receptor ACE2^16^. Present studies have indicated that the intercellular cell adhesion molecule-1 (ICAM1) was upregulated after IAV infection^28^, and ICAM1 has been reported as a receptor for RSV^29^, the interaction between IAV and RSV required further investigation. Among the positive interactions, IAV pre-infection showed the strongest promoting interactions on RSV infection. We performed confocal microscopy and observed more cell fusion (syncytia) in IAV and RSV coinfections than RSV mono-infections (data not shown), suggesting that IAV enhanced RSV infection by promoting the cell-to-cell transmission of RSV^30^. A recent study also observed a higher proportion of RSV-positive cells in IAV and RSV coinfections than RSV mono-infections^31^, while they and other studies showed the RSV titers were decreased in IAV and RSV coinfections than RSV mono-infections in the advanced stage of infection^32,33^, which was likely due to the depletion of cellular resources, resulting less RSV progeny released outside of the cells.

Among the negative interactions, both IAV and RSV negatively interacted with HPIV family. Intriguingly, the negative interactions between IAV and HPIV, or RSV and HPIV retained in the clinical analysis involving all-age cohorts^11,34^, implying these virus interactions between IAV or RSV and HPIV were independent of age. The inhibition against subsequent virus infection was likely due to the host immune response triggered by the initial virus infection. We also found the inhibition against HPIV3 by RSV was significantly attenuated in IFNAR^-/-^ cells compared with that in WT A549 cells (from 90.32-fold to 1.46-fold). Similarly, the inhibition against HPIV3 by IAV was also attenuated in IFNAR^-/-^ A549 cells (from 161.61-fold to 128.68-fold). However, even in the absence of IFN, IAV could still restrict HPIV3 infection, likely due to competing for the same host resources in coinfection.

People are curious about whether the directional virus-virus interactions, both positive and negative, among respiratory viruses could affect epidemiological dynamics. Our study found the strongest positive interactions among the HPIV family, which correspondingly manifested in a high level of synchronicity in their monthly prevalence time series. Similarly, a strong positive correlation existed between IAV and RSV, with their monthly prevalence time series exhibiting overlapping peaks of activity between Dec. and Feb. pre-COVID. In contrast, the negative correlations between viruses, such as IAV and HPIV3, correspond to staggered patterns in their monthly prevalence time series. Remarkably, in our cohorts, IAV infection cases dropped to zero from Mar. 2020 to Dec. 2021. Accordingly, RSV, positively correlated with IAV, and HPIV3, negatively correlated with IAV, also underwent significant changes. For instance, following the reduction of IAV, coinfections of RSV with other respiratory viruses significantly decreased (from 8.90% to 2.51%, 3.55-fold decreased). This suggests that the positive effect of influenza virus on RSV influences the coinfection ecology involving RSV. Similarly, coinfections among the HPIV family intensified following the reduction of IAV (e.g., the coinfection rate between HPIV2 and HPIV1 or HPIV3 increased from 76.3% to 96.6%). These findings indicate that influenza virus plays a pivotal role in the respiratory virus network, influencing the interactions and dynamics among other viruses.

In summary, our study suggests that virus interactions at molecular level could alter epidemiological dynamics and disease outcomes. When two positive correlated viruses co-circulate, more attention should be paid to virus coinfections and associated risk of severe disease. Our findings also highlight the pivotal role of influenza viruses in the respiratory virus interaction network, particularly in the post-pandemic era. As influenza viruses reclaim their niche in the respiratory ecology, this resurgence intensifies their epidemiological linkages and fosters an environment conducive to increased coinfections.

## Data Availability

All data produced in the present study are available upon reasonable request to the authors.

## Methods

### Study population and dataset

This study encompassed a cohort of 59,654 patients admitted to the Hubei Provincial Women and Children’s Hospital between January 2018 and December 2021. The patients who were hospitalized suffering from acute respiratory infection (ARI) aged no less than 18 years were enrolled. For each enrolled patient, throat swabs were collected and viral antigens were detected by using DHI seven respiratory virus detection reagents (immunofluorescence method) from DIAGNOSTIC HYBRIDS, INC. The seven respiratory viruses under investigation include influenza A virus (IAV), influenza B virus (IBV), respiratory syncytial virus (RSV), adenovirus (ADV), and human parainfluenza virus (HPIV) family, comprising HPIV1, HPIV2, and HPIV3. This comprehensive dataset is a reliable source for understanding the epidemiological patterns of respiratory illnesses, offering insights into common community-acquired respiratory virus infections within a sizable urban population.

Clinical outcomes of children aged 0-4 years were identified from electronic health records by using the hospital’s clinical data warehouse. We excluded the cases with congenital disease, including physical impairments (such as fracture), congenital heart disease, developmental delay of neonatal. For a comparable number of cases between mono-infection and coinfection, the cohort for clinical feature analysis was formed by randomly selected 237 (44-50 in each of the five indicated months) Flu mono-infection patients, randomly selected 242 (46-50 in each of the five indicated months) RSV mono-infection patients and all 46 coinfection patients. The multivariable logistic regression model was adjusted for the following variables: age, sex, admission temperature, bronchitis, pneumonia, hospitalization stays, and several laboratory parameters at presentation, including procalcitonin level, white blood cell count, lymphocyte count, hs-CRP level. As some patients did not have all laboratory studies of interest collected as part of clinical care, data are presented for only the patients in whom these were available. Patient medications including the anti-influenza virus drug, Oseltamivir were extracted from medication reconciliation fields in the electronic medical record.

The study was approved by the Medical Ethical Committee of the Maternal and Child Health Hospital of Hubei Province (2021-IEC-XM042), which waived the requirement for informed consent.

### Spearman correlation analysis

Spearman’s rank correlation coefficients were calculated and analyzed using a corrplot in R version 4.0.2 to assess the correlation between pairs of virus infection prevalences. The analysis focused on each virus pair’s monthly prevalence time series, with significance thresholds set at *Spearman’s rank correlation coefficients > 0.406 and **Spearman’s rank correlation coefficients > 0.521. Coefficients surpassing these thresholds were considered statistically significant.

### Cells and viruses

The A549, MDCK, and Vero-E6 cell lines were obtained from ATCC. The IFNAR^-/-^ A549 cell line, a generous contribution from Professor Ying Zhu at Wuhan University, and the Hep-2 cell line, a generous contribution from Professor Zishu Pan at Wuhan University, were used in the experiments. All cell lines were cultured in Dulbecco’s modified Eagle’s medium (DMEM; Gibco) supplemented with 10% fetal bovine serum (FBS) and incubated at 37 °C in a 5% CO2 environment.

The influenza A virus, specifically the A/WSN/33 virus, was generated using reverse genetics, following established procedures^35^. The influenza B virus (Yamagata lineage), Adenovirus (VR-1), and human parainfluenza virus 2 (VR-92) were obtained from the China Center for Type Culture Collection (CCTCC) and appropriately stored. Human respiratory syncytial virus A2 strain (VR-1540) was obtained from Professor Zishu Pan at Wuhan University. Human parainfluenza virus 3 (HPIV3) was obtained from Professor Mingzhou Chen at Wuhan University. All viruses were aliquoted and preserved at −80 °C until needed.

### Virus infection and IFN treatment

For virus infection, cells were washed with PBS and were subsequently incubated with IAV, IBV, and RSV at MOI of 0.1, or ADV, HPIV2, and HPIV3 at 0.14 TCID_50_, using the infection medium (DMEM supplemented with 2% FBS, 1% penicillin/streptomycin) at 37 °C in 5% CO_2_. IAV and IBV which were exposed for 1 h, and other respiratory viruses were exposed for 2 h. Subsequently, cells were washed twice with PBS and then incubated in the infection medium at 37°C in 5% CO_2_ for 24 h, except for ADV, which was incubated for 48 h.

For sequential coinfection, target cells seeded in 24-well plates were initially washed with PBS. Subsequently, cells were incubated with the primary virus (Virus A) for 12 h. At 12 h post-infection, cells underwent another wash with PBS and were then incubated with the second set of viruses (Virus B) for 24 h, except for ADV, which was incubated for 48 h. At 36 h or 60 h post-infection, 100 μl of supernatants were lysed with 400 μl TRIzol LS (Invitrogen^TM^,10296010). The cells were washed with PBS and then lysed with 500 μl TRIzol (Invitrogen^TM^,15596018).

For simultaneous coinfection, target cells in 24-well plates were initially washed with PBS and subsequently incubated with the two viruses in a final volume of 0.2 mL for 2 h, except for IAV and IBV coinfection, which was carried out for 1 h. After this incubation, cells were washed with PBS two times and were then incubated in the infection medium at 37 °C in 5% CO_2_ for 24 h, except for ADV, which was incubated for 48 h. Mono-infection with the same titer served as a control. At 24 h or 48 h post-infection, the cells were washed with PBS and then lysed with 500 μl TRIzol.

### Extraction of virus genome and Real-time quantitative polymerase chain reaction (qPCR)

The virus genome was extracted using virus DNA/RNA reagents 2.0 (Vazyme, RM401-01) and nucleic acid extraction instruments (Vazyme, VNP-32P). For RNA virus genome detection, DNAs were enzymatically digested by DNase I (Vazyme, EN401) for 20 min, and the DNase I activity was terminated with EDTA (Beyotime). Purified RNAs were subjected to reverse transcription using oligo dT and random primer (Takara, RR037A). Subsequently, the resulting cDNAs were quantified using Hieff qPCR SYBR Green Master Mix (Yeason). DNAs for ADV were directly quantified using Hieff qPCR SYBR Green Master Mix. The virus genome levels of the specified genes were quantified through quantitative PCR (qPCR). Thermal cycling was conducted in a 384-well reaction plate (ThermoFisher, 4343814). All virus genome levels in cells were normalized to the β-actin level in the same cell, while the virus genome levels in the supernatants were normalized to the negative control.

### Flow cytometry

Flow cytometry analysis was conducted to quantify virus infectivity. For IAV and RSV coinfection, A549 cells, seeded in 6-well plates, were washed with PBS and then incubated with IAV (MOI = 0.01) for 12 h. Following this, at 12 h post-infection, cells were washed with PBS and were subsequently incubated with RSV (MOI = 1) for an additional 12 h. At 24 h post-infection, cells were fixed in freshly prepared 4% (w/v) paraformaldehyde, permeabilized with 0.1% (v/v) Triton X-100, blocked with 1% BSA, and then stained with anti-influenza virus-NP antibody (provided by Professor Ningshao Xia), and anti-respiratory syncytial virus-F antibody (Fitzgerald, 10-R25H), followed by an APC-conjugated anti-rabbit secondary antibody or a FITC-conjugated anti-mouse secondary antibody. Samples were acquired using the Beckman Cytoflex flow cytometer and analyzed with FlowJo software. Uninfected cells were run in parallel for background subtraction. For IAV and HPIV3 coinfection, the procedures were consistent, except cells were stained with anti-human parainfluenza virus-F antibody (Fitzgerald, 10C-CR3016M1) followed by a FITC-conjugated anti-mouse secondary antibody.

### Immunofluorescence microscopy

For IAV and RSV infection, A549 cells were seeded on 12-well glass slides and allowed to adhere for a minimum of 24 h before use in experiments. Subsequently, A549 cells were infected with RSV (MOI = 0.1) for 24 h, with or without 12 h of IAV pre-infection (MOI = 0.1). After virus infection, cells were fixed in freshly prepared 4% (w/v) paraformaldehyde, permeabilized in 0.3% (v/v) Triton X-100 for 5 min, and blocked with 1 % BSA for 1 h. Subsequently, cells were immunostained with mouse primary antibodies against RSV F (Fitzgerald, 10-R25H) and rabbit primary antibody against IAV NP for 1 h at room temperature. Alexa Fluor dye-conjugated secondary antibodies (Alexa Fluor M488, Invitrogen; Alexa Fluor R568, Invitrogen) and DAPI (Beyotime, C1002) were used for 45 min at room temperature in the dark. After washing with PBS, cell-seeded glasses were mounted on slides with an antifade mounting medium and images were captured using a Zeiss Laser Scanning Microscope (LSM) 880 confocal microscope (Zeiss). For HPIV3 infection, the procedures were consistent, except cells were immunostained with mouse primary antibodies against HPIV3 F (Fitzgerald, 10C-CR3016M1).

### Mice

All animal experiments were performed in an ABSL-2 facility. Female BALB/c mice, 6-7 weeks old, were obtained from Charles River Laboratories. For sequential coinfection, mice were initially intranasally infected with either PBS or 1000 PFU of IAV. Subsequently, both groups were intranasally infected with 3 × 10^5^ FFU of RSV at 12 hours post-IAV infection. After another 48 hours, mice were sacrificed to determine viral loads and histological assays. The Animal Care and Use Committee of Wuhan University approved all animal experiments.

### Histology analysis

Lung tissue from infected mice was dissected on hour 60, fixed, and subjected to a standard Hematoxylin-Eosin staining (H&E) procedure. The slides were scanned and analyzed by the Wuhan Sci-Meds company. Representative images from three mice in each group were presented. The pathological score includes (i) alveolar septum thickening; (ii) recruitment and infiltration of inflammatory immune cells; (iii) hemorrhage. Each feature assessed was assigned a score of: 0, no substantial findings; 1, minimal; 2, mild; 3, moderate; 4, moderate to severe; 5, marked or severe.

### Statistical analysis

If not specified otherwise, an unpaired Student’s t-test was employed for two-group comparisons. Significance levels were denoted as follows: *p-value < 0.05, **p-value < 0.01, ***p-value < 0.001, and ****p-value < 0.0001. Unless otherwise indicated, error bars represent mean values and standard deviations from at least three biological experiments.

## Author contributions

K.X. conceived the project and designed the experiments. J. X., C.W., SM.L., and X.L. collected and analyzed clinical data. J.X. and C.W. provided clinical data and information and contributed to data interpretation. SM.L., X.L., YL.Z., X.N., and L.Q. coordinated virus coinfection experiments and conducted data analysis. L.Q. and SM.L. performed immunofluorescence, confocal microscopy, and flow cytometry experiments. SM.L. and X.L. performed animal infection experiments and evaluated the immunofluorescence and histopathological studies. C.Z., YA.Z., S.L., C.S., Z.P., M.C., and K.L. provided technical support and materials. K.X., SM.L., YL.Z., and Y.L. wrote the manuscript with input from all the other authors. All authors approved the final manuscript.

